# Evaluation of two red cell inclusion staining methods for assessing spleen function among sickle cell disease patients in North-East Nigeria

**DOI:** 10.1101/2023.01.12.23284472

**Authors:** Adama I Ladu, Ngamariju A Satumari, Aisha M Abba, Fatima A Abulfathi, Caroline Jeffery, Adekunle Adekile, Imelda Bates

## Abstract

**Introduction:** The loss of splenic function is associated with an increased risk of infection in sickle cell disease (SCD); however, spleen function is rarely documented among SCD patients in Africa, due partly to the non-availability of sophisticated techniques such as scintigraphy. Methods of assessing splenic function which may be achievable in resource-poor settings include counting red blood cells (RBC) containing Howell Jolly Bodies (HJB) and RBC containing silver-staining (argyrophilic) inclusions (AI) using a light microscope. We evaluated the presence of HJB - and AI - containing RBC as markers of splenic dysfunction among SCD patients in Nigeria.

**Methods:** We prospectively enrolled children and adults with SCD in steady state attending outpatient clinics at a tertiary hospital in North-East Nigeria. The percentages of HJB- and AI-containing red cells were estimated from peripheral blood smears and compared to normal controls.

**Results:** There were 182 SCD patients and 102 healthy controls. Both AI- and HJB-containing red cells could be easily identified in the participants blood smears. SCD patients had a significantly higher proportion of red cells containing HJB (1.5%; IQR 0.7% - 3.1%) compared to controls (0.3%; IQR 0.1% - 0.5%) (*P* = 0.0001). The AI red cell counts were also higher among the SCD patients (47.4%; IQR 34.5% - 66.0%) than the control group (7.1%; IQR 5.1% - 8.7%) (*P* = 0.0001). The intra-observer reliability for assessment of HJB-(R = 0.92; R^2^ = 0.86) and AI-containing red cells (R = 0.90; R^2^ = 0.82) was high. The estimated intra-observer agreement was better with the HJB count method (95% limits of agreement, −4.5 to 4.3; *P* = 0.579).

**Conclusion:** We have demonstrated the utility of light microscopy in the assessment of red cells containing - HJB and AI inclusions as indices of splenic dysfunction in Nigerian SCD patients. These methods can be easily applied in the routine evaluation and care of patients with SCD to identify those at high risk of infection and initiate appropriate preventive measures.

## Introduction

The spleen plays an important role in protection against infection and individuals with loss of spleen function are at an increased risk of infection with encapsulated microorganisms and systemic spread (1). In sickle cell disease (SCD), the spleen undergoes a sequence of pathological changes that ultimately result in loss of splenic function (2), and this accounts for most of the early morbidity and mortality associated with this disorder (3). The tests often used to assess spleen function are based on scintigraphy, haematological, and immunological techniques (4, 5). The scintigraphy method is the gold standard and assesses the ability of the spleen to filter blood of abnormal cells and particles. However, this method is expensive and involves injecting radio-labelled substances into patients (4, 5). The haematological methods reflect the inability of the spleen to phagocytose poorly deformable red blood cells (RBCs) or those containing inclusions. Splenic dysfunction is usually evaluated on the basis of increased numbers of such abnormal RBCs in circulation. The presence of Howell-Jolly bodies (HJB) is one such example; percentages of HJB red cells can be estimated from peripheral blood smears using the classical May-Grunwald Giemsa stain (MGG) (6), by flow cytometry (7) or imaging flow cytometry methods (8). In patients with diminished splenic function, the membrane of the red blood cells contain surface indentations referred to as ‘pits’ when viewed under an interference phase-contrast microscope (5). The pitted red cells count method represents a sensitive technique for evaluating splenic function(9); however, it requires specialised equipment and personnel. Another method of evaluating spleen function is counting red cells containing silver-stained (argyrophilic) inclusions (AI). The silver stain was originally developed to study the nucleolar organizer region of chromosomes and to evaluate their function (10, 11). The technique was first employed to assess splenic function by a group of investigators in the USA, following their observation that red cells from SCD patients and splenectomised individuals contained large numbers of AI (12). The AI count method is non-invasive and requires only a light microscope.

Spleen function has not been well studied in SCD patients residing in Africa (13), the region with more than two thirds of the global burden of SCD (14). This is mainly because most of the methods mentioned above are not readily available in most low-income settings of Africa as evidenced by the scarcity of data in the literature. The HJB and AI methods may be suitable but have not been widely used in Africa; both tests require only a light microscope and therefore are feasible in Nigeria and most low-to-middle-income countries. They can be used to facilitate the measurement of splenic function in SCD patients where resources for spleen scintigraphy and interference contrast microscopy are absent. Improvement in spleen function has been reported following chronic hyper-transfusion (15, 16) and following hydroxyurea therapy (17, 18). The splenic filtration function was preserved after three years in a third of the patients following hydroxyurea therapy. Therefore, assessment of splenic function among SCD patients in low-income countries can play a vital role in identifying those patients who can benefit from such therapy. The objective of the current study was to assess splenic function by comparing the frequencies of red cells containing HJB and AI in SCD patients with those of normal controls. We also evaluated which of the method was associated with the least intra-observer variation. If splenic function could be assessed in this manner, these techniques might be utilized for early identification of SCD patients at risk of developing severe infection because of reduced or absent splenic function, who may benefit from more intensive infection prevention measures.

## Methods

### Study participants

This was a prospective cross-sectional study conducted amongst children and adults with SCD in steady state between September 2020 and November 202; all patients who presented to the outpatient paediatric and adult haematology clinics of the University of Maiduguri Teaching Hospital (UMTH) during the study period were invited to participate. We excluded patients who had received blood transfusion in the last 3 to 4 months as this could interfere with the peripheral blood counting for AI and HJB red cells, as well as those with other acute or chronic illnesses that might interfere with spleen function. All SCD patients also underwent abdominal ultrasonography to document the presence or absence of the spleen. Apparently-healthy individuals consisting of medical students and children of hospital staff, with no recent history of fever or evidence of any underlying haematological disorder were enrolled as controls. The controls were of similar age range to the SCD participants.

### Ethics

Institutional review board approval was obtained from both UMTH (REC reference number: 20/606) and Liverpool School of Tropical Medicine (REC reference number: 20-010). Written and verbal consent was obtained from all the participants.

### Laboratory analysis

### Blood smears

Two thin blood smears, each for AI and HJB red cells estimation, were made from blood samples collected by venepuncture from every participant within 4 hours of collection and allowed to air dry. Smears for AI red cells were fixed for 3 minutes in a solution made by diluting 150 ml ethanol to 450 ml formalin (37% formaldehyde), washed in water, and allowed to dry thoroughly before proceeding to staining. Smears for HJB red cells were fixed in absolute methanol for 1 minute and allowed to air dry before staining with May Grunwald Giemsa (MGG) as described below. All fixed smears were stored in slide boxes if staining was not performed immediately.

### Method for AI count using silver stain

The silver stain method was based on the technique described previously (12), with some modifications (S1 Appendix 1). A pilot study to assess and refine the test performance was performed using samples from normal controls and SCD patients who were not participants in our study. We conducted a pre-test run to check the effect of staining duration, temperature, the constitution of eosin, and pre-treatment of slides with potassium iodide, on optimal staining conditions. As a result, the staining duration was reduced to 20 minutes in the final study protocol, as a longer duration was associated with over-stained slides that could not be interpreted (S1 Fig). The eosin was constituted using an aqueous solution as this provided better counterstaining ability, compared to an alcohol-based formulation. The reduction of slides with potassium iodide did not produce any added effect; therefore, this step was not included in the final protocol. Furthermore, two different brands of silver nitrate salts (50%; Honeywell Fluka, J3090 and FujiFilm Wako 196-00831) and Gelatin powder (500g: Sigma Aldrich Lot no BCBZ6147 and Riedel De Haen AG, Seelze Hannover, 18807) were tested and both gave the same staining quality. Finally, not more than 3 ml of working solution was prepared per staining session, as the stain began to deteriorate after about 3 minutes of preparation (S2-S4 Figs). The slides were stained using silver stain at 38^0^C for 20 minutes as described (S1 Appendix 1). All stained slides were mounted, as this improved the clarity of microscopy and reduced the effect of reflection observed with unmounted slides.

### Method for quantitative HJB count using May-Gruenwald Giemsa (MGG) stain

To obtain a uniform quality of staining, the slides were stained simultaneously in batches using a staining tank containing freshly made May-Gruenwald working solution for 5 minutes and washed in 2 to 3 changes of water. The slides were next immersed in Giemsa stain for 10 minutes before washing in 2 to 3 changes of tap water. The slides were allowed to dry upright as this prevented residues from forming on the slides. Any residual water on the slides was wiped off with dry gauze, which also improved the clarity of the film.

### Microscopy

Microscopy was performed using a light microscope (Leica DM 750, equipped with a ICC50 E colour camera) at 400x and 1,000x magnification. Five hundred RBCs for the AI and 400 RBCs for the HJB were counted per smear. The AI and HJB counts were expressed as percentages of the total red cells counted. Two blood smears each for AI and HJB red cells from each patient were examined separately for the percentages of cells of interest; the mean of the two counts observed was then calculated. To remove intra-observer variation and improve the precision of the microscopy results, all counts were performed by a single person (AIL). The reader had undergone prior training and quality checks during the pilot phase of the project.

### Statistical analysis

The data were entered into Excel spread sheet for cleaning and sorting and exported into Statistical Package for the Social Sciences (SPSS) (version 25; SPSS, Chicago, IL, USA) and MedCalc ® v.20.114 (MedCalc Statistical Software Ltd, Ostend, Belgium) for analysis. Categorical data were summarized using frequency and proportions while continuous data were expressed as means, median and interquartile range as appropriate. For the AI and HJB methods, the intra-observer reliability and agreement between two separate measures performed for an individual sample were assessed. Firstly, scatter plots were drawn to assess the relationship between the two separate readings. The Pearson’s correlation analysis was used to assess the intra-observer reliability for the two separate counts obtained using each method. The Bland Altman analysis was used to measure the limit of agreement between the two separate readings (19); individual values were expressed relative to the geometric mean of the first and second counts for that sample. A non-parametric test was used to compare results between SCD patients and controls. The level of significance was set at the two-tailed P-value <0.05.

## RESULTS

### General characteristics

Blood smears for identification of red cells containing AI and HJB were obtained from 182 SCD patients (175 Hb SS, 5 Hb SC, and 2 Hb SB thal) and 102 controls (93 Hb AA, 7 Hb AS, and 2 Hb AC). The median ages for the SCD group and controls were 11 years (range 1 - 45 years) and 12.0 years (range 1 - 32 years) respectively.

### Identification of argyrophilic inclusion-positive (AI) and HJB red cells

Both AI- and HJB-containing red cells could be easily identified in the participants; representative smears for AI (Fig.1) and HJB are shown below (Fig. 2). The AI inclusions were spherical, oval or irregular in shape, of variable sizes and occurred either as single or multiple inclusions within a red cell (Fig. 1). The HJB appeared as single, spherical inclusion often situated close to the cell periphery (Fig. 2)

**Fig. 1:**
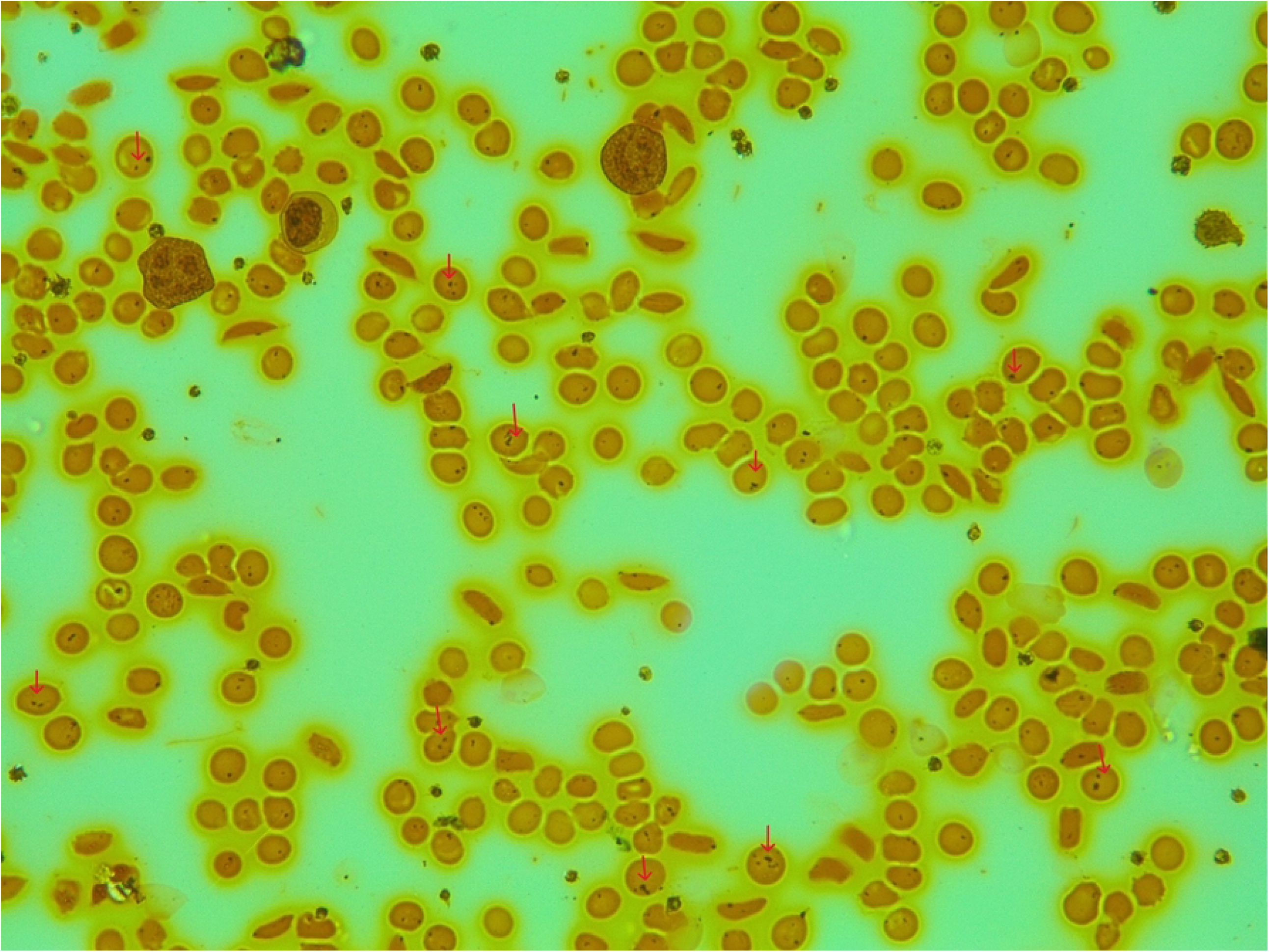
Image for argyrophilic inclusion (AI) containing red cells. Silver-stained blood smear for a 16-year-old SCD patient showing argyrophilic inclusions (arrows). The inclusions varied in size and numbers. Several sickled red cells also noted (x 40).

**Fig. 2:**
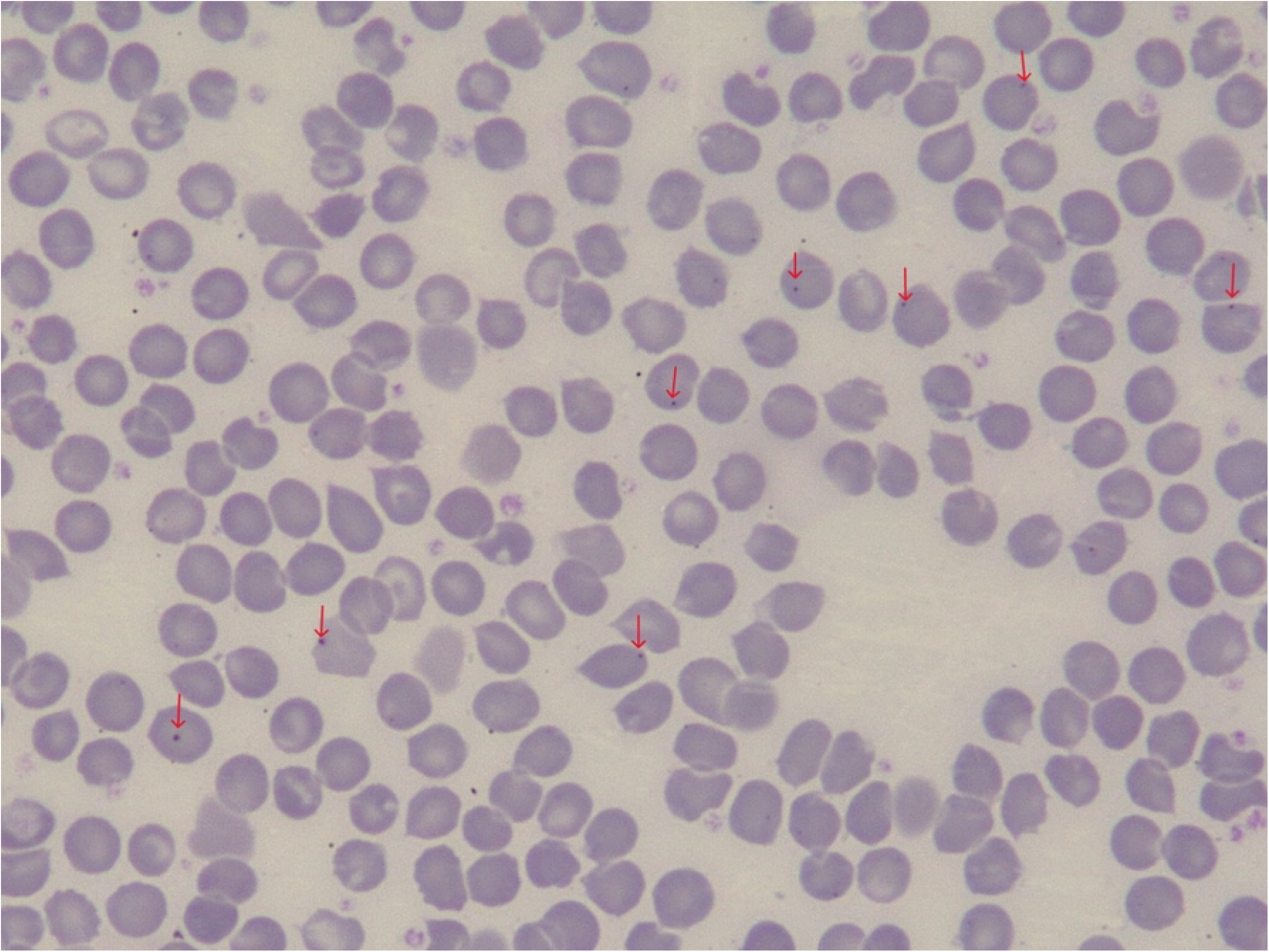
Image for Howell Jolly body (HJB) containing red cells. May-Grunwald Giemsa (MGG) stained blood smear for a 23-year-old SCD patient showing HJB inclusions (arrows) (x40)

### Comparison of AI and HJB red cells count among the study population

The distribution of HJB and AI red cells across the study population is shown in table 1. The median percentage of HJB red cells in the SCD group (1.5%; IQR 0.7% - 3.1%) was greater than those of the control group (0.3%; IQR 0.1% - 0.5%) (*P* = 0.000). Similarly, the median percentage of AI red cells in the SCD group (47.4%; IQR 34.5% - 66.0%) was significantly higher than those of the control group (7.1%; IQR 5.1% - 8.7%) (*P* = 0.000) (Mann-Whitney U test). Analysing the results according to Hb phenotype (Kruskal Wallis test), within the SCD group, the HJB and AI red cell counts tended to be higher in the HbSS population compared to the other compound heterozygotes, however, the results were not statistically significant either for the HJB (*P* = 0.883) or AI counts (*P* = 0.534). The small number of the latter group (5 Hb SC and 2 Hb SB Thal) may have affected the power to detect any statistical significance among the SCD group. Among the control group, the median percentage of AI red cells for those with Hb AA was not significantly different from those with Hb AS and Hb AC (*P* = 0.296). Similarly, the frequency of HJB red cells in the Hb AA controls was comparable to those of Hb AS and Hb AC controls (*P* = 0.946).

**Table 1:**

**Title: Distribution of AI and HJB red cell counts across study population**

**Footnote: AI: Argyrophilic inclusions; HJB: Howell Jolly Bodies; RBC red blood cells; Hb: haemoglobin. * 75^th^ centile is not generated because of only 2 data in the group**

### Comparison of frequency of red cell inclusions using the silver stain and MGG stain method

We compared the range of results obtained for red cell inclusions using both HJB and AI methods. It was observed that the results were always higher with the silver stain method for AI red cells. Among the SCD group, the AI red cell counts ranged from 34.5% to 66.0% and HJB counts ranged from 0.7% to 3.1%. Among the controls, the AI red cells count ranged from 5.1% to 8.7% whereas the HJB counts ranged from 0.1% to 0.5%.

### Comparison of frequency of AI and HJB red cells based on the presence or absence of spleen on ultrasonography among the SCD patients

The SCD patients were divided into two groups based on the presence (n=101) or absence (n=71) of the spleen on abdominal ultrasonography. The frequencies of AI and HJB red cells were compared between the two groups (using Mann Whitney U test). The median percentage of AI red cells was significantly higher in the group without visible spleens (56.0%; IQR 38% - 73%) compared to those whose spleens were visible on ultrasonography (44.9%; IQR 29% - 61%) (*P* = 0.008). Similarly, the median percentage of HJB red cells in the group without visible spleens on ultrasonography was higher (2.2%; IQR 1.5% - 5.1%) compared to those whose spleens were visible (0.9%; IQR 0.6% - 2.0%) (*P* = 0.0001)

### Intra-observer reliability between counts obtained for the AI and HJB method

Scatter plots showing the relationship between two separate readings obtained from the same sample for both the AI and HJB methods are shown below (Figs: 3 and 4). The Pearson’s correlation (R) for the AI and HJB counts was 0.90 (R^2^ = 0.82) and 0.92 (R^2^ = 0.86) respectively.

**Fig. 3:**
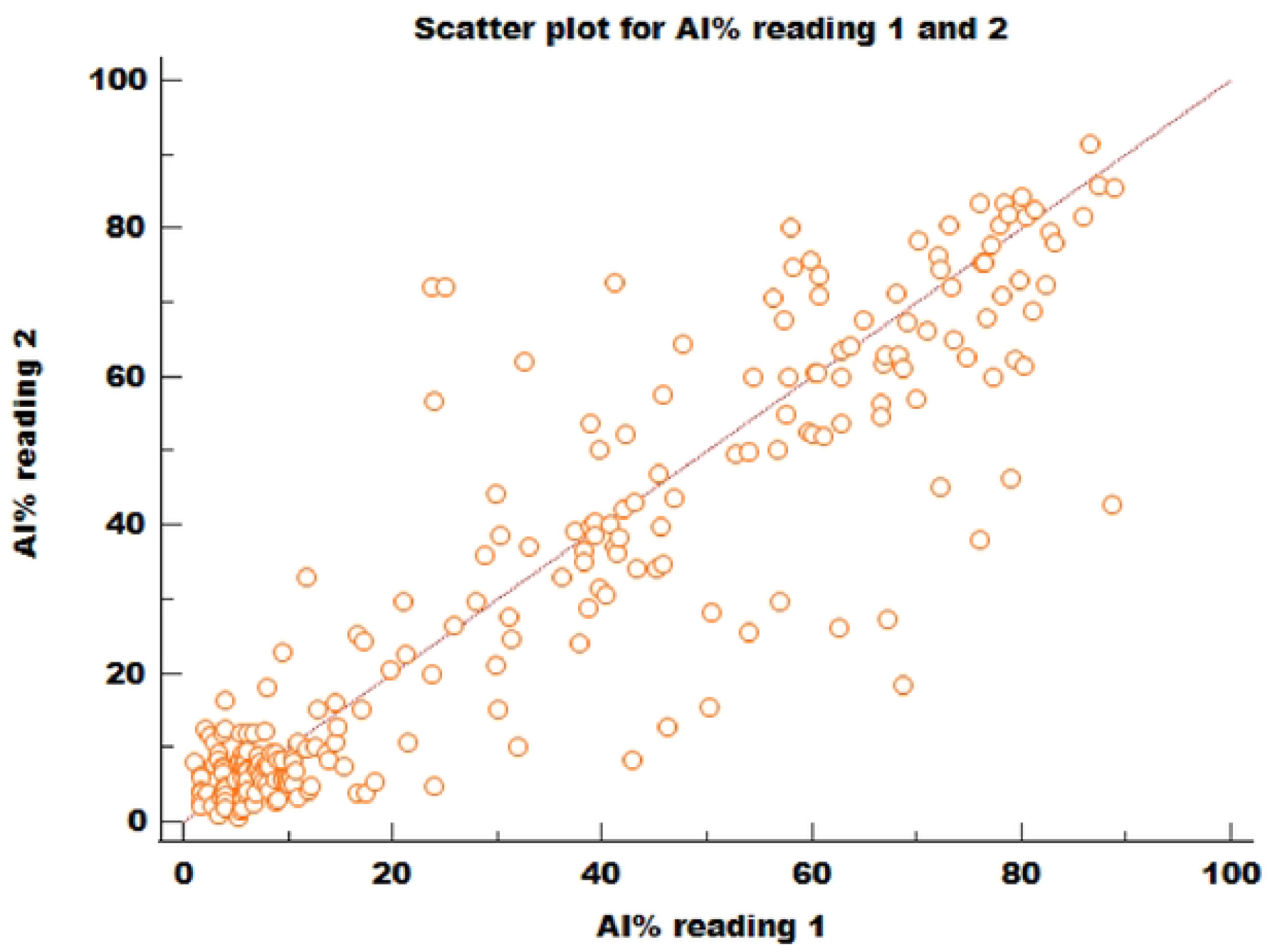
Plot for paired AI% red cells reading. Scatter plot showing the correlation between two different sets of counts for argyrophilic inclusion (AI) red cells (R = 0.90; R^2^ = 0.82). A total of 233 paired samples (both SCD and controls) were analysed.

**Fig. 4:**
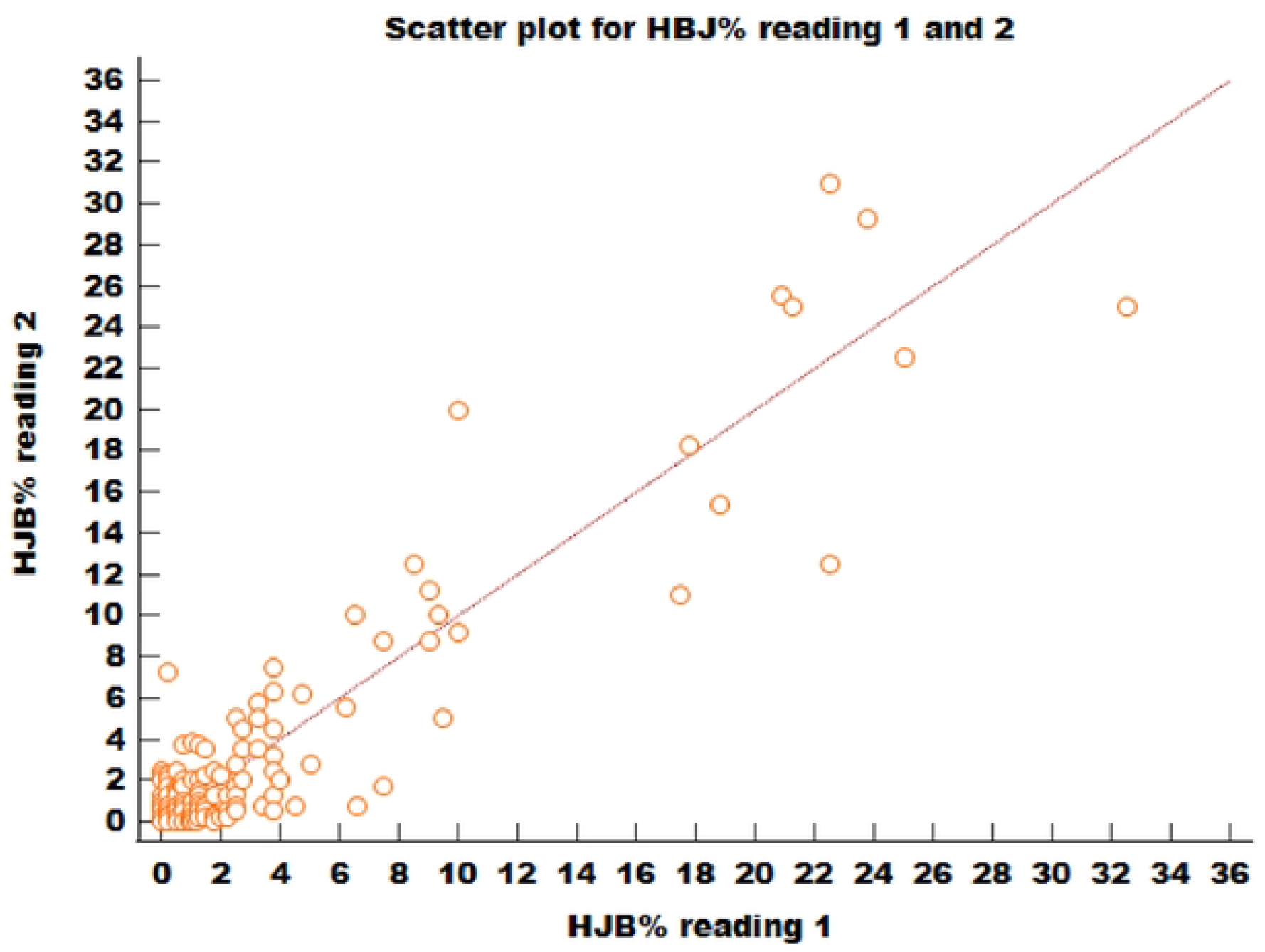
Plot for paired HJB% red cells reading. Scatter plot showing correlation between two different sets of counts for Howell-Jolly body (HJB) red cells (R = 0.92; R^2^ = 0.86). A total of 174 paired samples (SCD patients only) were analysed.

### Intra-observer agreement between counts obtained for the AI and HJB method

The mean difference between the two sets of HJB readings was low (difference −0.1; 95% limits of agreement −4.5 - 4.3) and non-significant (*P* =0.579) (Fig.5). Although, the points appeared clustered towards the lower end, they were scattered relatively evenly above and below the mean line of equality. The results of AI count showed a weak agreement between the two sets of counts when all the data sets were analysed (i.e., both SCD and control data). The mean difference between the two sets of readings was high (difference 1.7; 95% limits of agreement −22.2 - 25.6) (*P* = 0.03) (Fig.6a). However, when only the data sets from the control population were analysed, there was good agreement between the two sets of reading evidenced by the significantly low mean difference (difference 0.34; 95% limits of agreement −9.9 – 10.6) (*P* = 0.526) (Fig 6b).

**Fig. 5:**
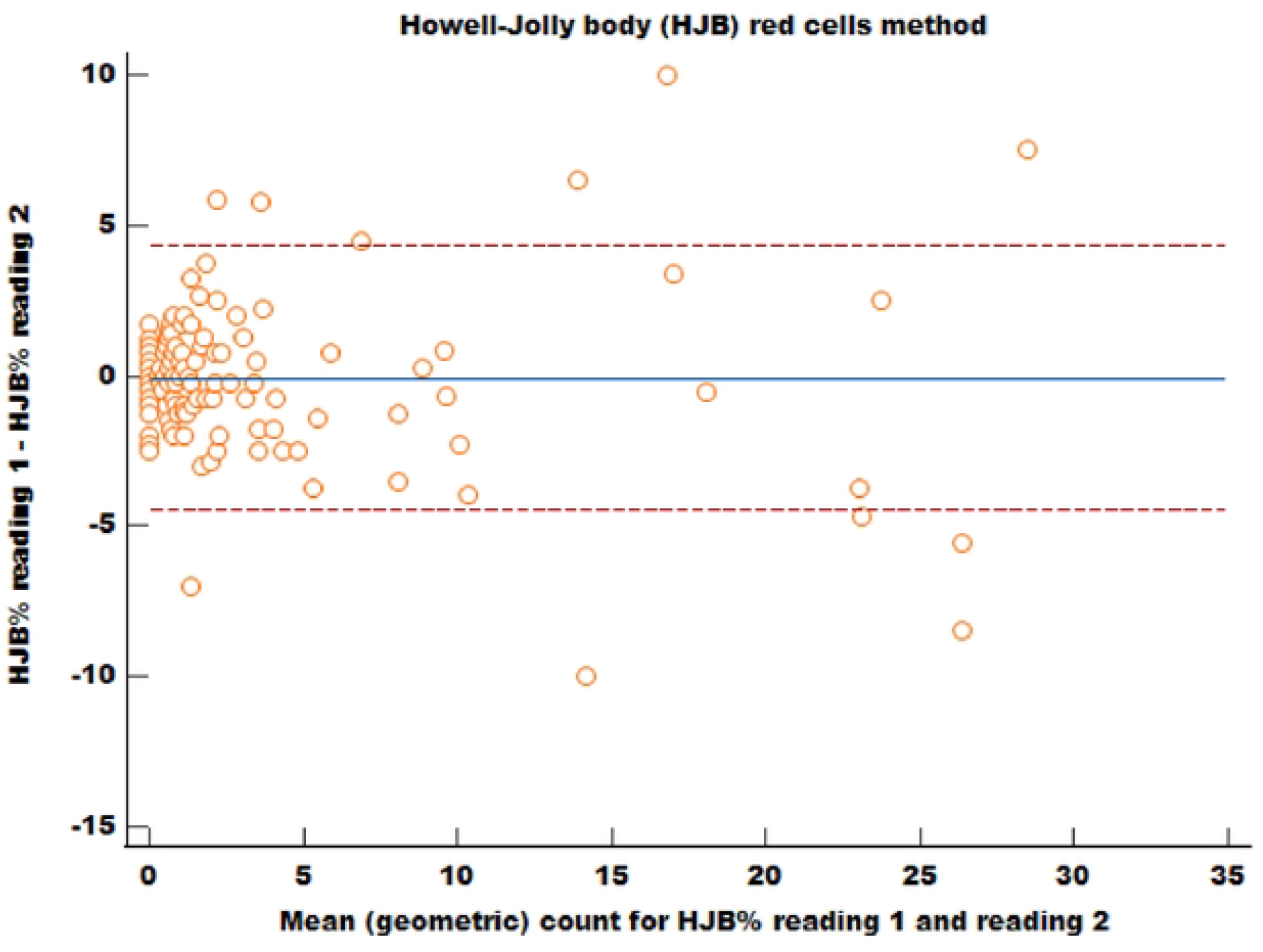
Howell-Jolly bodies (HJB) red cell method. Bland & Altman plot to demonstrate the limit of agreement between 2 different sets of counts for HJB red cells (174 paired samples from the SCD patients were analysed). X axis: Mean (geometric) count between the first and second HJB reading. Y axis: difference between the first and second HJB reading for an individual sample. The horizontal middle line represents the mean difference between the first and second reading and the dashed lines represent the 95% upper and lower limits of agreement.

**Fig. 6:**
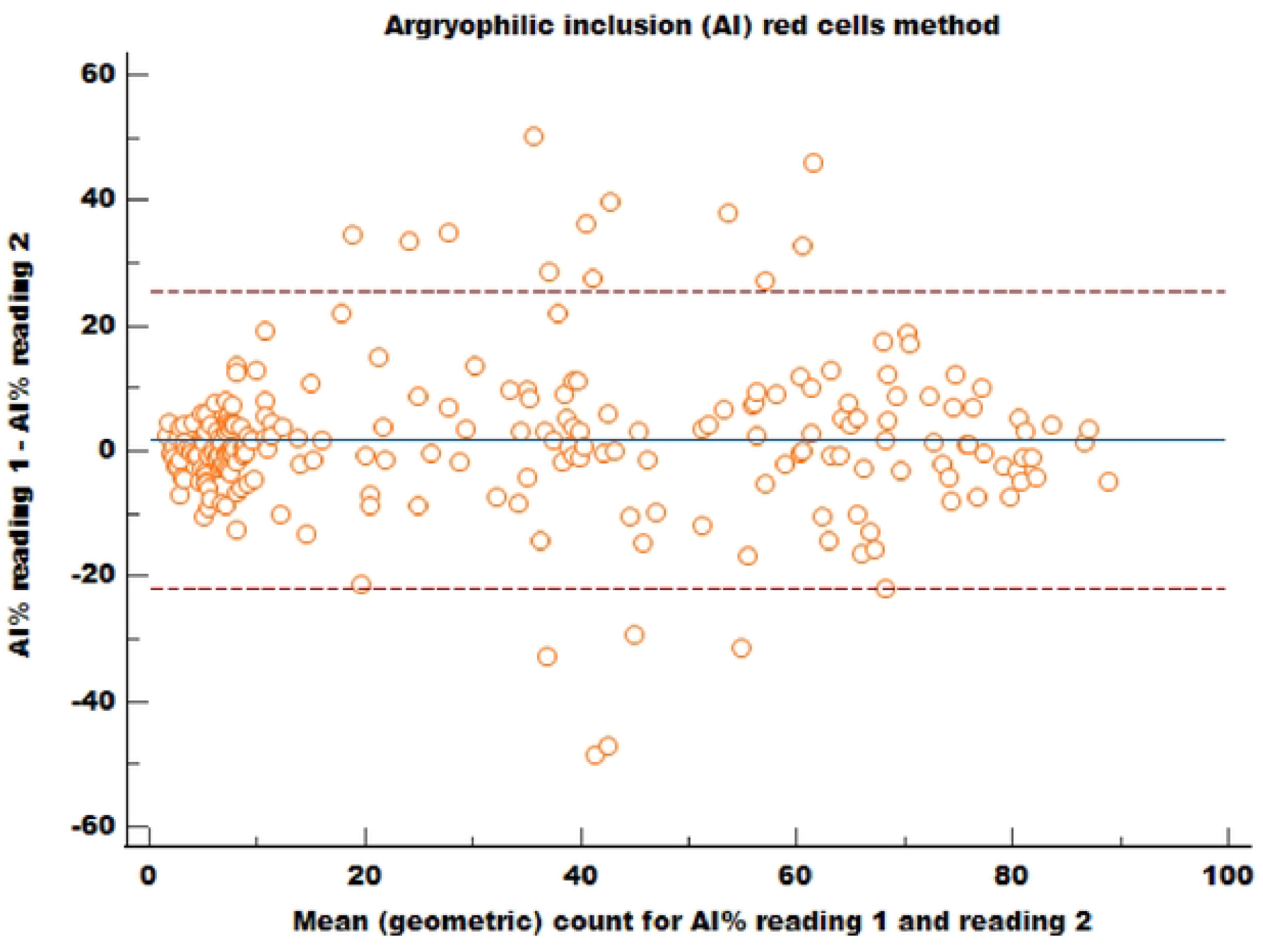
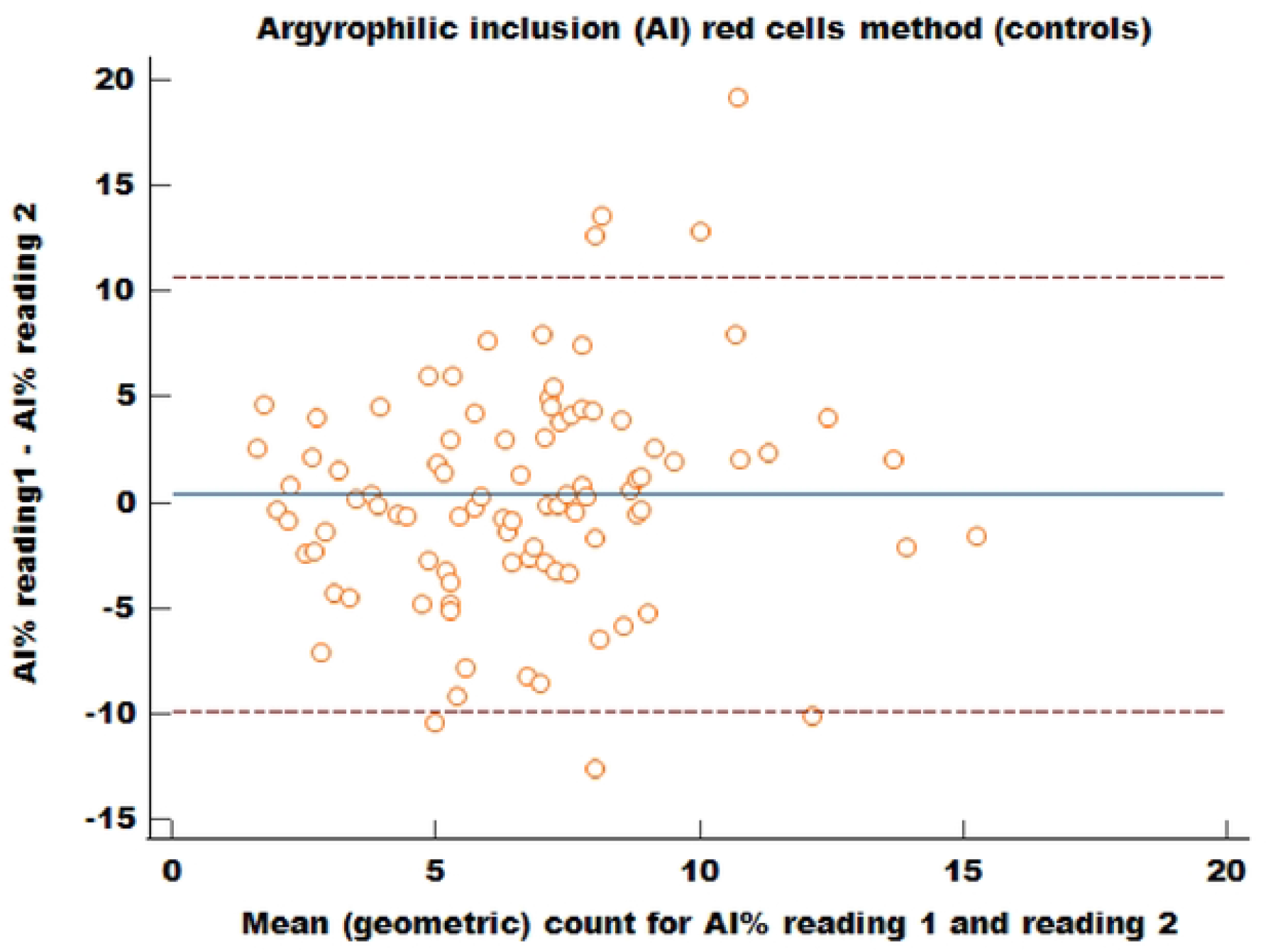
Argyrophilic inclusions (AI) red cell method. Bland & Altman plot to demonstrate the limit of agreement between 2 different sets of counts for the AI red cells. X axis: Mean (geometric) count between the first and second AI reading. Y axis: difference between the first and second AI reading. The horizontal middle line represents the mean difference between the first and second reading and the dashed lines represent the 95% upper and lower limits of agreement. (a) (upper figure) represents data set for both SCD and controls (n=233) (b) (lower figure) represents data sets for only controls (n=10*2*)

## Discussion

Spleen function is usually evaluated by the spleen’s ability to remove abnormal cells from circulation. SCD patients develop progressive splenic dysfunction and quantification of red cells containing two types of inclusions - HJB and AI, was employed in the current study to evaluate their splenic function. The silver stain method used in the current study to evaluate AI-containing red cells as a marker of spleen function was based on that first described by Tham et al in the United States (12). The exact nature of the AI inclusions is unclear; however, studies have demonstrated the presence of vacuoles and inclusions among circulating red cells (9, 20, 21). The inclusions were increased in individuals without spleens or in those with abnormalities of erythropoiesis. They were more abundant in splenectomised persons with concurrent hematologic disorders such as thalassemia and haemolytic anaemia. Their increased presence in abnormalities of erythropoiesis may be readily explained by the larger amount of product of haemolysis to be disposed of in these conditions. Their increase however in splenectomised individuals, with otherwise normal erythropoiesis, suggests an intact spleen is required for their elimination. In one study, the authors noted inclusions in 54.3% of red cells from 20 splenectomised individuals, using interference phase-contrast microscopy (21). These inclusions are probably equivalent to the AI inclusions observed in the current study using the silver stain, in close agreement with our observed mean value of about 50% for AI red cells. The authors noted the inclusions neither stained nor looked like conventional red cell inclusions, such as HJB, Heinz bodies or siderotic bodies. Morphologically, the inclusions were suggestive of haemoglobin degeneration. They concluded that mature normal red cells continually form inclusions which may reflect the degradative cellular process consequent upon cell aging. Kent and colleagues also observed, by electron microscopy, the presence of autophagic vacuoles in mature red cells and reticulocytes; the inclusions contained a variety of materials including haemoglobin and altered cytoplasmic organelles such as ribosomes, mitochondria, and smooth membranes (20). The inclusions appeared to be instrumental in the disposal of those organelles or other materials not required by the fully mature red cells. The increased presence of these inclusions in splenectomised subjects indicates that, although developing human red cells are capable of eliminating their digestive residues, the presence of the spleen is required to fulfil this task adequately.

The use of MGG to demonstrate HJB in red cells is a long-established method of evaluating splenic dysfunction (3, 22, 23). HJB are small (approximately 1 micron) red cell DNA inclusions that result from cytogenetic damage. Normally, HJB-containing red cells are formed at low frequency and are quickly removed by an intact spleen, thus their presence in peripheral smears is an indirect evidence of splenic dysfunction (23, 24). Our comparison of results obtained using HJB and AI inclusions as markers of splenic dysfunction showed the AI in red cells were always higher than the HJB counts. This indicates the two tests identify different intra-cellular structures. While the silver stain picks up all intracellular argyrophilic particles, MGG stains only HJB, which are usually formed at low frequency. The argyrophilic inclusions varied in size, shape, and number within the red cells, whereas the HJB always appeared as a single inclusion. The higher AI count may also be related to the increased erythropoiesis seen in haemolytic disorders like SCD. Kent al found increased red cells with inclusions in patients with haematological disorders and reticulocytosis but intact spleens; a high proportion of the reticulocytes contained inclusions (20). We also observed in our study that the reticulocytes percentage correlated with the AI count (rho = 0.23; P= 0.002) but not with the HJB count (rho = 0.14; P = 0.07). Whatever their nature, it appears the AI are found in increased numbers in SCD patients compared to HJB; the forma although more sensitive in picking up red cells inclusions, may be influenced by both splenic dysfunction and haemolysis related abnormalities.

### Comparison between SCD patients and controls

To our knowledge, this is the first detailed study of splenic function among SCD patients in an African setting. Our study found significant differences in the frequencies of AI and HJB red cells between the SCD patients and normal controls, which indicates the presence of splenic dysfunction in the former. The percentage of AI red cells in our SCD population was significantly higher than those of the controls; such a finding using the AI red cells has been reported in the US (12), though the study only included 9 SCD patients and 45 controls. However, the results obtained for the AI red cells in our SCD patients (IQR 34.5% - 66.0%) and controls (IQR 5.1% - 8.7%) were higher than the data from the SCD patients (11.8% - 52.7%) and controls (0.3% - 3.0%) in the American study (12). The SCD patients in the present study, demonstrated a higher frequency of HJB red cells compared to the controls. The range of HJB red cells obtained among our SCD population was higher (IQR 0.7% - 3.1%) than results obtained among SCD patients (n = 12; age range 5 months to 39 years) in the United States (HJB% range 0 - 1.1%) (25) and among children (n = 20; age range 5 years to 22 years) with SCD in Brazil (HJB% range 0 - 1.4%) (26). The small sample size from both studies compared to our sample population (n=182) may account for the differences compared to our results.

### Comparison of AI and HJB red cells in SCD patients with spleen present or absent on scan

Both the AI and HJB red cell counts from SCD patients with autosplenectomy were higher than patients whose spleens were visualized on ultrasound. This is not unexpected as the spleen is the site of removal of red cell inclusions, therefore patients with autosplenectomy are likely to have higher numbers of inclusions within their red cells than those with intact spleens. However, despite the presence of the spleens in some of the SCD patients, the frequency of red cell inclusions was still higher than in normal controls. This suggests the presence of functional hyposplenism among SCD patients, whereby despite having the spleen present, it may not be effectively eliminating the inclusions (24). Although, the effect of age on the AI and HJB count among the SCD patients was not addressed in the current study in order to identify the onset of splenic dysfunction, this is being reported in another paper focusing on the clinical implications of the current study findings.

### Reliability and agreement analysis for the HJB and AI methods

To evaluate which of the method would give the least intra-observer variation and be useful for the longitudinal follow up of our patients, we evaluated the intra-observer reliability and agreement of both methods. Our data demonstrated a good correlation between the two sets of reading obtained with the AI test (R = 0.90; R^2^ = 0.82), in keeping with an earlier report (r=0.736; *P* <0.001) (12). While correlation measures the linear relationship between two sets of measurement, agreement on the other hand assesses the equality of the individual values between two sets of measurement (27). Since a perfect positive correlation is not proof of equal responses between two testing occasions (28), we tested results obtained for both the AI and HJB methods for agreement using the Bland Altman analysis (19). A good agreement was observed for the AI counts among the controls (*P*=0.499), but at higher counts obtained among the SCD patients, the AI method did not produce such a good agreement between two readings taken on different occasions. This finding reflects the character of the AI test; the difference between two separate readings taken on the same sample is related to the value of the measurement, larger measurements among the SCD patients imply a larger average error between both readings and hence the significant difference between these counts (*P*=0.03).

Our data also showed a good limit of agreement for counts obtained using the HJB method (95% CI, −4.5 - 4.3; *P* = 0.579). Several investigators have shown the HJB count can be reliably used to assess spleen function. Corazza et al found a significant correlation between HJB red cell counts obtained by the classical MGG and the pitted red cells counts (P < 0.0001); they noted that pitted red cells count above 8% was always associated with increasing HJB red cells count (6). Serial measurements of HJB red cells using the manual estimation were reliably used to monitor spleen function before and after bone marrow transplantation in SCD patients over a period of 15 years (29, 30). In a similar, but small study, the frequency of HJB red cells declined progressively following bone marrow transplantation, and together with scintigraphy, both techniques were used to monitor spleen function in these patients (31). Also, a robust correlation between the MGG method and a newly-developed imaging flow cytometry method for evaluating HJB has been demonstrated (8). A few studies however have reported a lack of correlation between the MGG HJB method with other methods of assessing spleen function (25, 26).

A prospective study of SCD patients (n=12) identified at birth in a screening programme in the USA, demonstrated that splenic dysfunction is an acquired defect occurring as early as five months of age; the onset of splenic dysfunction documented by serial splenic scintigraphy correlated with the appearance of HJB in peripheral smears (22). This suggests that the presence of HJB can be used as a reliable indicator of the onset of splenic dysfunction. Our study indicates that counting of HJB red cells by the MGG method can be used to demonstrate splenic dysfunction in SCD. Compared to other currently available methods of assessing spleen function, such as spleen scintigraphy, pit red cells counts and flow cytometry, this method provides a simple and reliable technique that can easily be used in most laboratories in the African settings. HJB was easy to perform, simpler than the AI count method and can be used to evaluate splenic function at regular intervals. This can allow for the longitudinal observation of splenic function among SCD patients to identify those that begin to show an increase in their HJB counts. This will be valuable in identifying early the onset of splenic dysfunction, which is associated with increased susceptibility to overwhelming infections. Such individuals can benefit from early medical intervention and preventative measures.

## Limitations

To determine estimates of diagnostic accuracy such as sensitivity and specificity, we would have compared our findings with those of a reference standard such as the spleen scintigraphy or pitted red cell count (32). Since these methods are not available in Nigeria, we have compared our findings to those in the published literature.

## Conclusions

Determination of red cells with AI and HJB enables the assessment of splenic function in SCD patients in low-resource settings. Both markers can serve as indicators of splenic dysfunction in SCD patients as reflected by the high percentages of circulating levels compared to controls. Their presence was easily demonstrable, and both showed good intra-observer reliability. Although the red cells inclusions were readily demonstrable using the AI method, the silver stain deteriorates rapidly. The HJB method is simpler and requires fewer reagents than the AI method. Moreover, the higher AI counts in patients with haemolytic anaemias such as SCD may be due to both the hematologic disturbance and abnormal splenic function, therefore, further validation in larger studies may be required for the AI method before the generalizability of its efficacy in assessing spleen function in SCD can be ascertained.

## Data Availability

The data underlying this article are available in the manuscript, figures, and tables. Further request can be addressed to the corresponding author

## Acknowledgement

The authors are grateful for the assistance of the staff of Histopathology and Haematology department of UMTH during data collection, sample preparation and microscopy analysis of this project.

## Authors’ contributions

Conceptualization: Adama Ladu, Imelda Bates

Formal analysis: Adama Ladu Investigation: Adama Ladu, Ngamariju A Satumari

Methodology: Adama Ladu, Ngamariju A Satumari, Aisha M Abba, Fatima A Abulfathi

Supervision: Imelda Bates, Caroline Jeffery, Adekunle Adekile

Writing – original draft: Adama ladu

Writing – review & editing: Adama Ladu, Ngamariju A Satumari, Aisha M Abba, Fatima A Abulfathi, Caroline Jeffery, Adekunle Adekile and Imelda Bates

## Supporting information

Additional supporting information may be found online in the Supporting Information section at the end of the article.

**S1 Fig:**

**Title: Effect of prolonged staining of slide**

**Silver-stained smear for SCD patient showing deeply stained red cells (arrows) making visualization of some of the argyrophilic inclusions difficult**.

**S2 Fig**

**Title: Silver stain at 1 minute**

**Image showing a preparation of silver stain at one minute. The preparation appears clear and colourless**

**S3 Fig**

**Title: Silver stain at 3 minutes**

**Image showing a preparation of silver stain three minutes following constitution. The preparation has begun to take a yellow colouration**

**S4 Fig**

**Title: Silver stain at 4 minutes**

**Image showing a preparation of silver stain four minutes following constitution. The preparation has turned golden yellow and continues deteriorating on further standing**

**S1 Appendix 1:**

**Title: Standard operating procedure for silver stain technique**

